# Comparing India’s second COVID wave with the first wave – a single-center experience

**DOI:** 10.1101/2021.06.03.21258009

**Authors:** Mayank Kapoor, Budha O Singh, Prasan Kumar Panda, Pathik Dhanger, Anant Kataria

## Abstract

**Background:** The COVID-19 pandemic has resurfaced in India in the form of a hard-hitting second wave. This study aims to compare the clinical profile of the first wave (April-June 2020) and the second wave (March-May 2021) of the severe acute respiratory syndrome coronavirus 2 (SARS-CoV-2) in a single tertiary care center of India. This will help prioritize the target population group and management strategies in the upcoming third wave if any.

**Methods:** In this retrospective observational study, we examined the demographic profile, symptoms at presentation, the severity of illness, baseline investigations, treatments received, underlying comorbidities, and outcomes of the COVID-19 patients belonging to the first (W1) and the second wave (W2) of the pandemic in India.

**Findings:** Among 106 patients in W1 and 104 patients in W2, the age group affected most was 37·1 (SD=16·9) years compared with 50.5 (SD=17.7) years respectively. The baseline oxygen saturation is lower in W2, being 84·0 (13·4) % compared with 91·9 (7·4) % in W1. 70.2 % of the cases belonged to the severe category in W2 compared to 37.5% in W1. W2 patients demonstrated higher transaminase levels [SGOT, 108.3 (99.3) v/s 54.6 (69.3); SGPT, 97.6 (82.3) v/s 58.7 (69.7)] with respect to W1. Similarly, the CT severity score for W2 [29.5 (6.7)] was higher than W1 [23·2 (11·5)][All P<0.05]. The proportion of patients requiring oxygen (81.8% v/s 11.2%), high flow nasal cannula (11.4% v/s 5.6%), non-invasive ventilation (41.2% v/s 1.5%), invasive ventilation (24.5% v/s 0.9%), as well as ICU/HDU admissions (56.4% v/s 12.0%) was higher for W2 as compared with W1. The measured case fatality rate varies from 29% for W2 to 9.6% for W1.

**Interpretation:** Higher age, oxygen requirement, ventilator requirement, ICU admission, and organ impairment are more prevalent in the admitted COVID-19 cases during the second wave that has hit India compared to the first wave and associated with more fatalities. Strategy for another wave should be planned accordingly.

**Key points:** *Question:* What are the differences between the clinical profile of the first wave (W1) and the second wave (W2) of the severe acute respiratory syndrome coronavirus 2 (SARS-CoV-2) in a single tertiary care center of India?

*Findings:* In this observational study among 106 patients in W1 (April-June 2020) and 104 patients in W2 (March-May 2021), there were higher proportion of increased age, oxygen requirement, ventilator requirement, ICU admission, and organ impairment in the admitted COVID-19 cases during the second wave.

*Meaning:* The second wave hits India badly than the first wave and associated with more fatalities.

## Introduction

The Coronavirus disease 2019 (COVID-19) pandemic has taken the whole of the world in its grip. After initial containment, the virus has re-emerged in many countries very strongly. An excellent example is India, where after initial control, the second wave has struck more fiercely.^1^ The first wave showed a peak in mid-September 2020, with a maximum of about 97,000 cases per day. By January of this year, the daily caseload had decreased to less than 10,000 a day. However, starting from February, India’s COVID-19 cases began spiking, rising exponentially to more than four lakh cases per day in April 2021. The daily number of deaths reported also rose from about a minimum of 70 to about 3900 per day. The new variant is termed a “double mutation” variant, referring to mutations in the virus’s spike proteins.^2,3^ The mortality rate for the disease in India was 1.1%.^4^ Many speculations regarding the age group affected, the oxygen requirement trend, and the associated complications of the second wave (W2) in comparison to the first wave (W1) are there.

This study explores what makes the second wave different from the first, which has led to such an alarming spike of cases in India. We analyzed the data from patients admitted to tertiary care dedicated COVID-19 hospital during April-June 2020 (W1) and March-May 2021 (W2) to find the differences between the two pandemic waves in India.

## Methods

### Study design and participants

The study was carried out at All India Institute of Medical Sciences (AIIMS), Rishikesh, a tertiary care center catering to North India’s population. We retrospectively searched the hospital database and analyzed the records of lab-confirmed COVID-19 patients of at least 18 years of age. Positivity obtained on reverse transcriptase-polymerase chain reaction (RT-PCR) SARS-CoV-2 assay of nasopharyngeal or oropharyngeal swab specimens defined COVID-19 positivity using SD Biosensor standard M nCoV real-time detection kit, Biorad CFX 96 real-time thermocycler, and Thermo flex 96 extractor machine. The biochemical investigations were performed according to the institutional treatment protocols and standards. The treatment decisions were solely clinician-based according to the protocols. At the time of admission, the patients were divided into different categories according to the severity of the disease. We followed the Ministry of Health and Family Welfare (MoHFW), India guidelines (Table 1).^5^

**Table 1:**
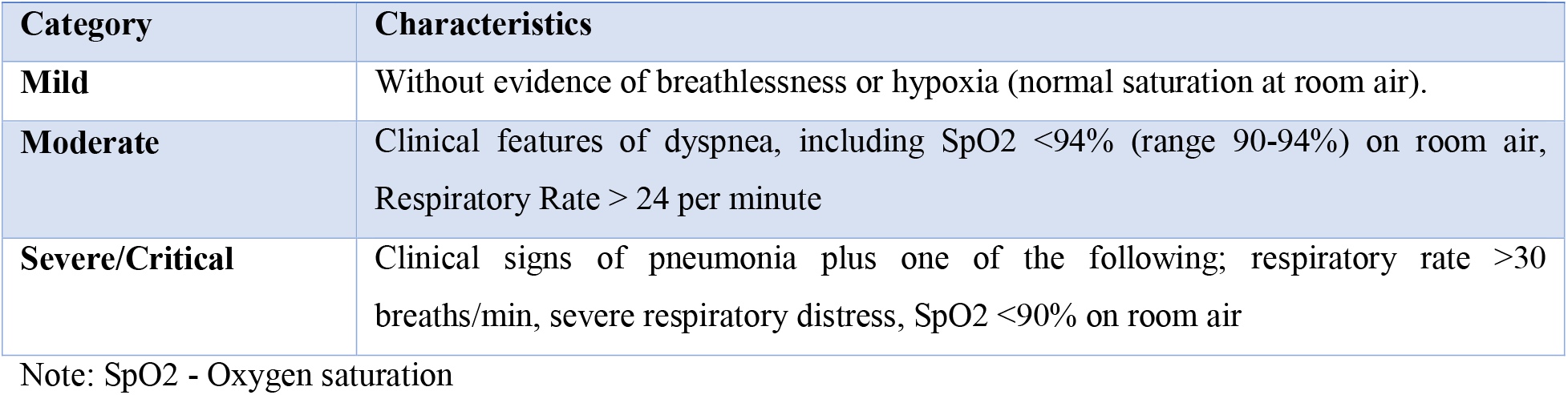
Ministry of Health and Family Welfare (MoHFW) COVID severity categorization^5^

| Category | Characteristics |
| --- | --- |
| **Mild** | Without evidence of breathlessness or hypoxia (normal saturation at room air). |
| **Moderate** | Clinical features of dyspnea, including SpO<sub>2</sub> <94% (range 90-94%) on room air, Respiratory Rate > 24 per minute |
| **Severe/Critical** | Clinical signs of pneumonia plus one of the following; respiratory rate >30 breaths/min, severe respiratory distress, SpO<sub>2</sub> <90% on room air |

Note: SpO<sub>2</sub> - Oxygen saturation

We compared the two groups based on their age, health risk factors (smoking status and presence of diabetes and hypertension), severity, and outcomes. We analyzed the number of patients requiring oxygen, non-invasive or invasive ventilation, ICU admissions, and the various treatments. Ethical approval was obtained (CTRI/2020/08/027169).

### Statistical analysis

Continuous data following normal distribution and homogeneity of variance was expressed as mean ± SD. We used χ ^2^ and Student’s t-tests to compare the demographic characteristics of individuals belonging to the two waves. Statistical analysis was done using SPSS version 25. P values <0.05 were considered significant.

## Results

Between April 2020 to June 2020 (W1), we analyzed records of 106 patients; and compared them with 104 patients from March 2021 to May 2021 (W2) (Fig. 1). The baseline mean oxygen saturation was found to be lower in W2 (Table 2). We had 70.2 % of the cases belonging to the severe category in W2 compared to 37.5% in W1. W2 patients demonstrated higher transaminase levels with respect to W1. Similarly, the CT severity score for W2 was also higher than W1. A higher proportion of patients required oxygen, high flow nasal cannula, non-invasive ventilation, invasive ventilation, and ICU/HDU admissions in W2 compared with W1 (Tables 3). COVID mortality was also higher in W2 than W1 (Tables 4).

**Fig. 1:**
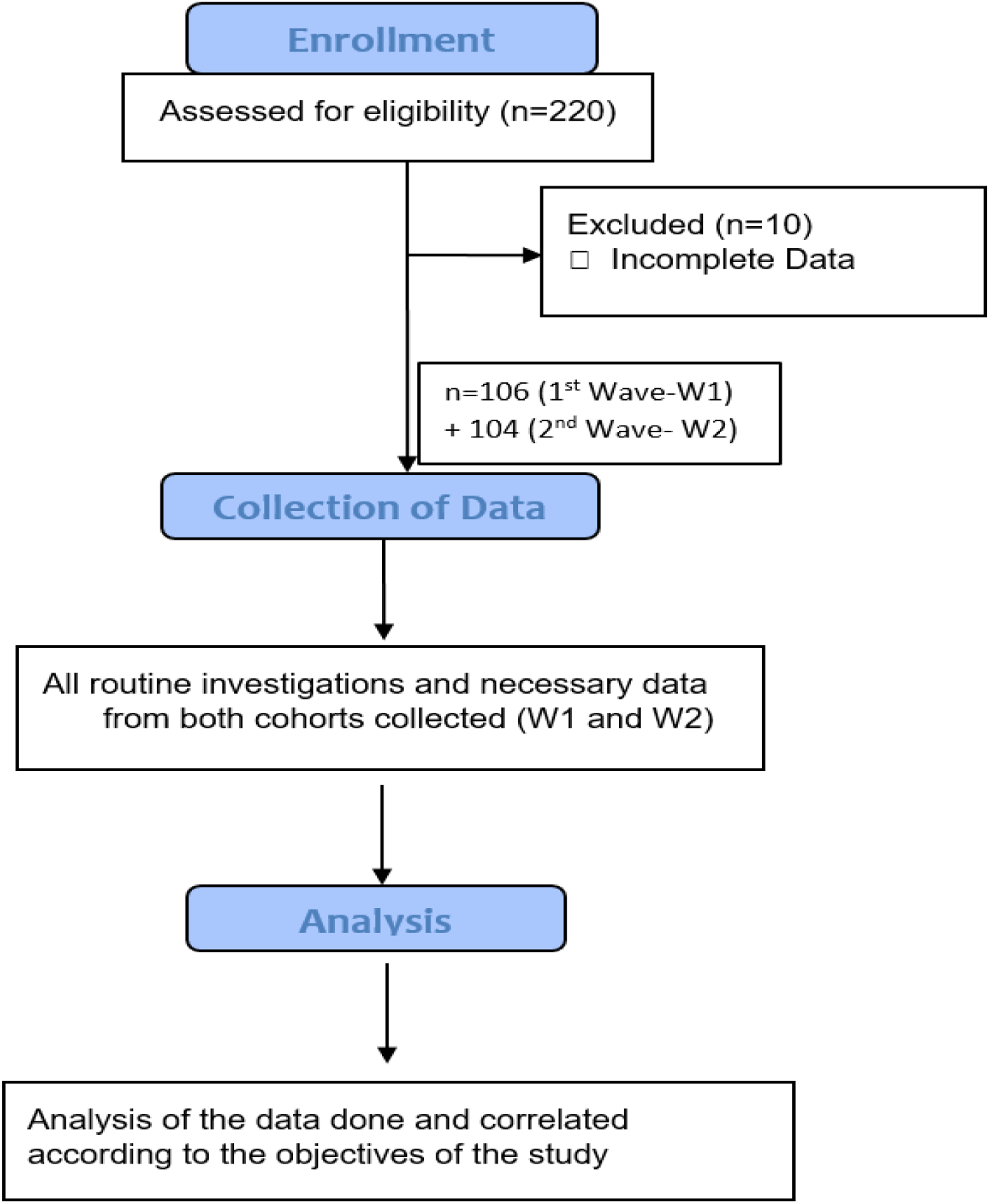
The study flow [W1 – First COVID-19 wave (April-June 2020); W2 – Second wave (March-May 2021)]

**Table 2:** Demographic characteristics of the study population during both COVID-19 waves

|  |  |  |  |
| --- | --- | --- | --- |
| Variable | Sub-group | First Wave (W1)<br>n=106 | Second Wave (W2)<br>n=104 |
| **Sex** | Female | 36 (34.4%) | 46 (44.3%) |
|  | Male | 70 (65.6%) | 58 (55.7%) |
| **Age, years<sup>§</sup>** |  | 37.1 (16.9) | 50.5 (17.7) |
| **Temperature (°F)<sup>§</sup>** |  | 98.5 (0.9) | 98.8 (1.2) |
| **Heart rate (per minute)<sup>§</sup>** |  | 91.0 (12.3) | 92.9 (14.5) |
| **Respiratory rate (per minute)<sup>§</sup>** |  | 20.0 (3.6) | 25.1 (4.7) |
| **SpO2 (%)<sup>§</sup>** |  | 91.9 (7.4) | 85.5 (13.4) |
| **Severity** | Mild | 42 (39.8%) | 15 (14%) |
|  | Moderate | 24 (22.6%) | 16 (15.8%) |
|  | Severe | 40 (37.5%) | 73 (70.2%) |
| **Symptoms** | Fever | 31 (29.3%) | 88 (84.5%) |
|  | Cough | 15 (14.2%) | 80 (77.5%) |
|  | Sore throat | 12 (11.1%) | 5 (5%) |
|  | Runny nose (Rhinorrhea) | 3 (3%) | 3 (3%) |
|  | Shortness of breath (Dyspnea) | 16 (15%) | 83 (80%) |
|  | Loss of smell / taste (Anosmia/Aguesia) | 1 (1%) | 4 (4%) |
|  | Fatigue / Malaise | 10 (9.5%) | 19 (18.6%) |
|  | Diarrhea | 12 (11.1%) | 5 (5%) |
| **Associations** | Hypertension | 13 (11.9%) | 31 (30.3%) |
|  | Diabetes Mellitus | 18 (17.6%) | 24 (22.8%) |
|  | Smoking | 5 (4.6%) | 1 (1%) |

Data are expressed as number (percentage) unless otherwise stated
<sup>§</sup>Data is expressed as mean (SD)

**Table 3:**
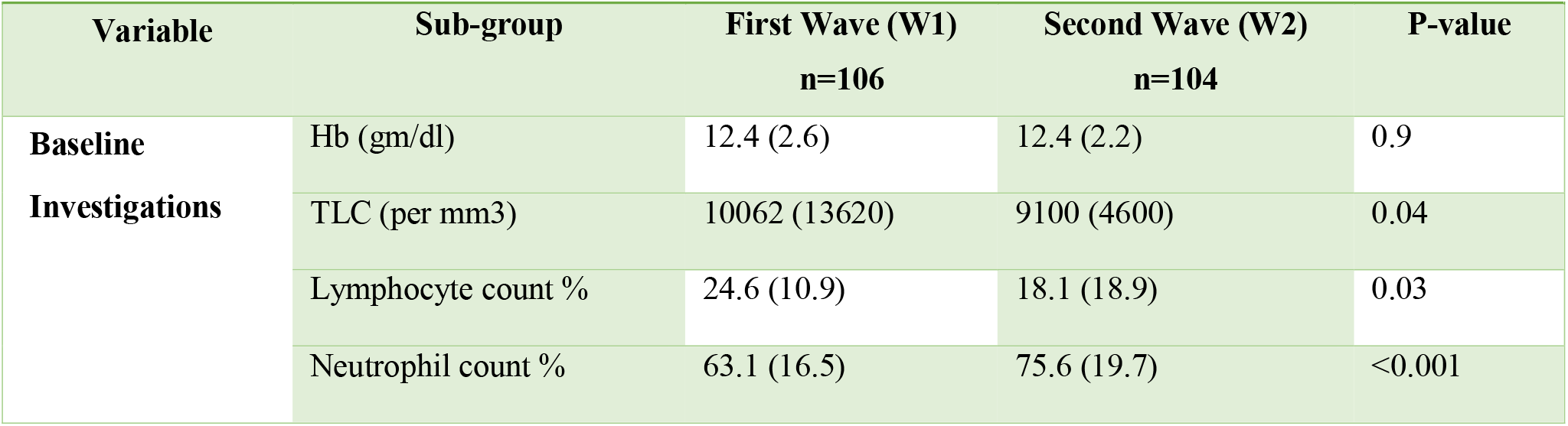
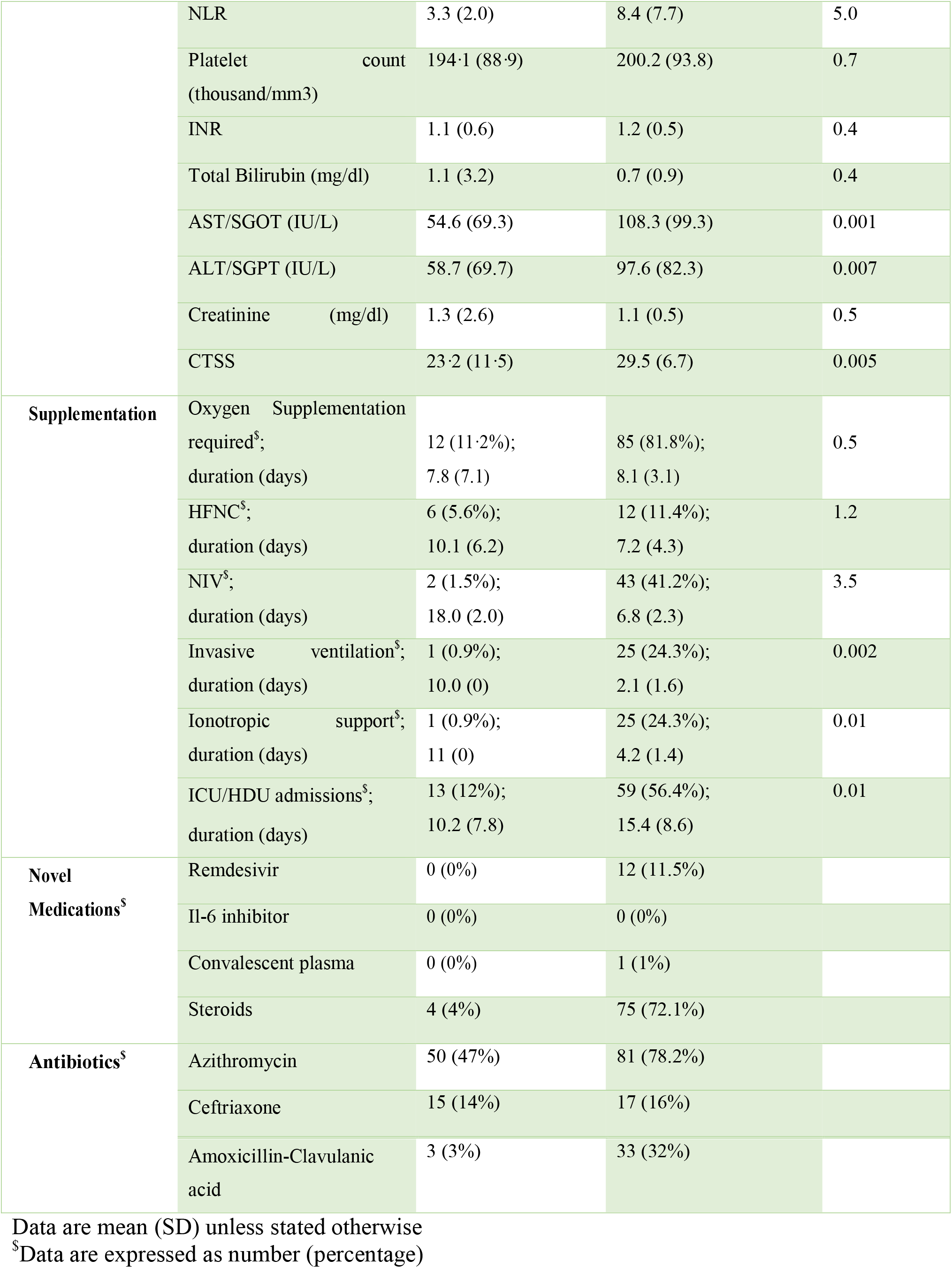
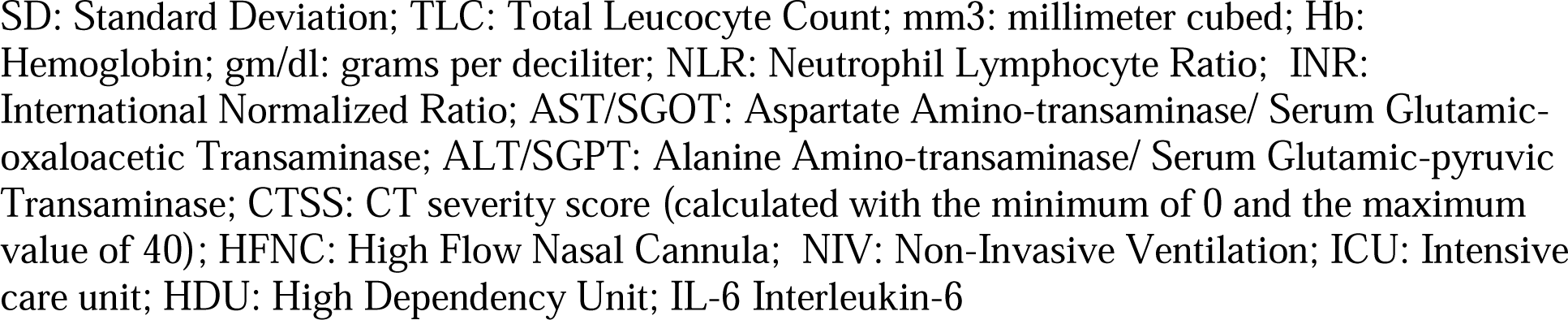
Routine investigations and treatment received during both COVID-19 waves

**Table 4:** Complications and outcomes of patients during both COVID-19 waves

| Variable | Sub-group | First Wave (W1) n=106 | Second Wave (W2) n=104 |
| --- | --- | --- | --- |
| **Complications** | Bacterial pneumonia | 1 (0.9%) | 9 (9%) |
|  | ARDS | 7 (6.4%) | 51 (48.7%) |
|  | Pleural effusion | 0 (0%) | 3 (2.5%) |
|  | MI | 2 (1.8%) | 3 (2.5%) |
|  | Pulmonary embolism | 0 (0%) | 0 (0%) |
|  | AKI | 3 (2.5%) | 19 (18%) |
|  | ALI | 7 (6.4%) | 13 (12.8%) |
|  | Hyperglycemia | 10 (9.6%) | 15 (14%) |
| **Outcome** | Discharge | 96 (90.4%) | 74 (71%) |
|  | Death | 10 (9.6%) | 30 (29%) |

Data are expressed as number (percentage)
ARDS: Acute Respiratory Distress Syndrome MI: Myocardial Infarction, AKI: Acute Kidney Injury, ALI: Acute Liver Injury

## Discussion

The COVID-19 pandemic, although contained in some parts of the world, is still re-emerging in many countries, especially this second wave. This has led to the re-introduction of lockdown in some countries.^6^ This shows that still, we are far away from altogether tackling this pandemic. The second wave of COVID hitting India is in many ways different from the first.

In this large-scale retrospective study, we have analyzed the major differences between COVID W1 and W2. In W2, patients are presenting with more dyspnea (80% v/s 15%) and hypoxia [Spo2 84·0 (13·4)% v/s 91·9 (7·4)%]. This has large logistics implications for developing countries like India. Hence, we recommend urgent commission of oxygen plants so that these upcoming oxygen demands can be met. 70.2% of the cases in W2 have the severe disease compared with 37.5% in W1; thus, ICU admissions are more in W2 (56.4%) than W1 (12.0%). Differences are most likely due to varying demographic structures and virus strain characteristics, among other factors. The novel coronavirus has differential dissemination; hence, certain regions have a higher disease burden than others for reasons not understood. After acquiring a vast sea of data from various trials, we also found that hospitals in W2 are utilizing various novel therapies like remdesivir (11.3% v/s 0%) and steroids (72.1% v/s 4%) in higher frequencies as compared with W1. However, this has associated complications too, like hyperglycemia (14% v/s 9.6%) and a dreaded infection rapidly rising in the immunocompromised COVID patients in India, Mucormycosis.^7^

The W2 has worse outcomes too. Incidence of acute respiratory distress syndrome (48.7% v/s 6.45), acute kidney injury (18% v/s 2.4%), acute liver injury (12.8% v/s 6.4%), and deaths (29% v/s 9.6%) is higher in W2. All these suggest that W2 is hitting India harder than W1; hence timely action is the need of the hour to accommodate the different demographic distribution of the second wave of COVID infection in India utilizing proper social distancing and sanitizing protocols.^8^ The higher levels of SGOT and SGPT values in W2 need to be studied further. In addition, frequent monitoring of routine blood investigations must be done to watch for renal/hepatic/other organ dysfunction, which may add to the list of poor prognostic factors in COVID-19.

Our study has limitations. We only analyzed the records of the hospitalized patients; hence, we could not take into account any non-hospitalized COVID-19 patient with mild disease.

Moreover, because of hospital bias, the proportion of severe cases is higher in W2 than W1. The primary reason for this being a change in the admission criteria from W1 to W2. In W1, the hospital had the policy of admitting all COVD-19 positive patients. However, with the rising number of cases and decreasing bed availability in W2, the hospital admits a predominating number of moderate and severe cases. In addition, the use of experimental therapies has appeared midway after the first wave in India, which is why their proportion of use is more in W2.

In conclusion, the second wave of COVID has hit India hard and can overwhelm India’s healthcare infrastructure. Hypoxia and organ dysfunctions are more frequently reported in the second wave population, leading to higher ICU admissions and deaths, further overburdening the situation. Our data could be used to inform India’s population about the severity of this second wave so that people understand the nature of the situation and follow all the COVID-19 appropriate behaviors more strictly. The lessons learned from this fight are essential to prevent a third wave from entering any country. We should be better prepared in terms of testing, vaccination, and boosting our critical care sector.

## Data Availability

It will be available by contacting the corresponding author.

## Contributors

MK contributed to the data collection, data analysis, and was involved in manuscript writing. BOS, PD, and AK contributed to the data collection and were involved in reviewing the draft. PKP gave the concept, interpreted analysis, critically reviewed the draft, and approved it for publication along with all authors.

## Data sharing

It will be made available to others as required upon requesting the corresponding author.

## Acknowledgment

None

